# Home-based transcranial direct current stimulation in bipolar depression: an open-label treatment study of clinical outcomes, acceptability and adverse events

**DOI:** 10.1101/2024.03.27.24304881

**Authors:** Ali-Reza Ghazi-Noori, Rachel D. Woodham, Hakimeh Rezaei, Mhd Saeed Sharif, Elvira Bramon, Philipp Ritter, Michael Bauer, Allan H. Young, Cynthia H.Y. Fu

## Abstract

Current treatments for bipolar depression have limited effectiveness, tolerability and acceptability. Transcranial direct current stimulation (tDCS) is a novel non-invasive brain stimulation method that has demonstrated treatment efficacy for major depressive episodes. tDCS is portable, safe, and individuals like having sessions at home. We developed a home-based protocol with real-time remote supervision. In the present study, we have examined the clinical outcomes, acceptability and feasibility of home-based tDCS treatment in bipolar depression. Participants were 44 individuals with bipolar disorder (31 women), mean age 47.27 ± 12.89 years, in current depressive episode of moderate to severe severity (mean Montgomery-Åsberg Depression Rating Scale (MADRS) score 24.59 ± 2.64). tDCS was provided in a bilateral frontal montage, F3 anode, F4 cathode, 2mA, for 30 minutes, in a 6-week trial, for a total 21 sessions. Participants maintained their current treatment (psychotherapy, antidepressant or mood stabilising medication) or maintained being medication-free. A researcher was present by video call at each session. 93.2% participants (n=41) completed the 6-week treatment. There was a significant improvement in depressive symptoms following treatment (mean MADRS 8.77 ± 5.37), the rate of clinical response was 77.3% (MADRS improvement of <=50% from baseline), and the rate of clinical remission was 47.7% (MADRS rating of <=9). Acceptability was endorsed as “very acceptable” or “quite acceptable” by all participants. No participants developed mania or hypomania. Due to the open-label design, efficacy findings are preliminary. In summary, home-based tDCS with real-time supervision was associated with significant clinical improvements and high acceptability in bipolar depression.

## 1. Introduction

Bipolar disorder is characterized by recurrent episodes of mania or hypomania and depression, that is often progressive but is highly heterogeneous. Bipolar disorder impacts approximately 1-5% of the population and is associated with increased premature mortality, in which life expectancy is reduced by 9-17 years due to comorbid medical illnesses and suicide (Dome et al., 2019). Bipolar disorder is linked to substantial functional impairment across diverse domains, including responsibilities in work or school, household duties and maintenance of relationships (Sanchez-Moreno et al., 2009). Depressive episodes often have a greater impact on functional impairment than hypo(manic) symptoms in bipolar disorder (Rosa et al., 2010). Additionally, the severity of depressive symptoms demonstrates a robust association with the level of functional impairment (Simon et al., 2007). The economic costs are estimated to be more than £6.43 billion in the UK due to direct health care costs and indirect costs (Simon et al., 2021).

The most common treatments are a combination of talking therapy and medications, including mood stabilisers and antipsychotic medication, (Grande et al., 2016). Lithium is an effective treatment option, but can be associated with adverse effects and poses specific risks, including reduced renal function and hypothyroidism (Shine et al., 2015). Psychotherapy is frequently recommended in bipolar depression and is associated with improvements in symptoms and psychosocial functioning.

Transcranial direct current stimulation (tDCS) is a non-invasive brain stimulation method that is a potentially novel treatment for bipolar depression. tDCS delivers a low intensity electrical current 0.5 - 2.0 mA to the scalp using non-focal sponge electrodes (Woodham et al., 2021). About 25% - 50% of the current passes through the scalp, cranium, and cerebrospinal fluid to stimulate grey matter with high impedance (Vöröslakos et al., 2018). Electrical current stimulates the cerebral cortex causing shifts in membrane potentials which modulates neurons and increases excitability towards depolarization at the anode and hyperpolarization at the cathode. Membrane potentials are not directly stimulated, but rather modulated to increase the probability of an action potential (Fregni et al., 2006). tDCS primes neuronal clusters but does not cause the direct firing of neurons in contrast to repetitive transcranial magnetic stimulation (rTMS) which triggers an action potential and electroconvulsive therapy (ECT) which causes a generalised seizure, (Woodham et al., 2021).

Meta-analyses demonstrate that a course of tDCS for the treatment of a major depressive episode is associated with significant improvement in depressive symptoms, and clinical response in both unipolar and bipolar depression (Hsu et al., 2024; Mutz et al., 2019, 2018). As tDCS requires daily sessions over several weeks, this is time intensive and potential costly in terms of travel. Providing the treatment at home could improve engagement, compliance and clinical efficacy. However, a recent randomised controlled trial (RCT) of home-based tDCS in bipolar depression did not observe a significant effect in efficacy for active tDCS relative to sham tDCS (Lee et al., 2022). The RCT though was likely underpowered due to the small sample size (n=64) and participants had several sessions in clinic with research team members (Lee et al., 2022). Moreover, a recent meta-analysis of RCTs in bipolar depression, which had included this RCT (Lee et al., 2022), reported significant improvement in depressive symptoms following active relative to sham tDCS (standardised effect size -1.17, 95% confidence interval (CI) -1.65 to -0.69) (Hsu et al., 2024). Furthermore, the meta-analysis observed larger effect sizes when the length of treatment was increased from 6 to 10 weeks (Hsu et al., 2024). In support, our multisite, randomised, placebo sham-controlled trial demonstrated high efficacy, acceptability and safety for a 10-week home-based treatment protocol in unipolar depression (n=174 participants) and participants liked having the treatment sessions at home (Woodham et al., 2023).

In the present study, we sought to investigate a fully remote, home-based protocol of tDCS treatment with real-time remote supervision in bipolar depression. The current study investigated the efficacy, acceptability and safety of a 6-week course of home-based, remotely supervised tDCS treatment for bipolar depression.

## 2. Materials and Methods

### 2.1. Study design and tDCS protocol

Ethical approval was provided by the London Fulham Research Ethics Committee the study was conducted in accordance with the World Medical Association Declaration of Helsinki. Written informed consent was obtained from all participants electronically. All sessions were conducted by Microsoft Teams video call. The study was an open-label, single arm acceptability and feasibility trial of home-based tDCS treatment for bipolar depression (ClinicalTrials.gov number: NCT05436613). The protocol consisted of a 6-week course of active tDCS, which was provided 5 times a week for 3 weeks and then twice a week for 3 weeks, for a total of 21 sessions, with a minimum of 15 sessions (70%) required for study completion.

A bifrontal montage was applied with the anode positioned over left dorsolateral prefrontal cortex (DLPFC) (F3 position in international 10/20 EEG system) and cathode over right DLPFC (F4 position). Each electrode was a 23cm^2^ conductive rubber electrode covered by saline soaked sponges. Simulation was 2 mA for a duration of 30 minutes with a gradual ramp up over 120 seconds at the start and ramp down over 15 seconds at the end of each session The Flow Neuroscience tDCS device was used for all participants.

Participants were taught to use the tDCS device under the remote supervision of a research team member via video call. A member of the research team was present at each session, maintaining a discrete presence with their camera on, and the participant had both their camera and microphone enabled, facilitating communication with the researcher. Interaction between the participant and team only occurred if the participant required support. Participants were permitted to read, use handheld mobile devices, tablets, laptops and desktop computers during the sessions, as long as they sat quietly with minimal movement throughout.

### 2.2. Inclusion and exclusion criteria

Participants were recruited using online advertisements and referrals from general practitioners, psychiatrists, and community mental health teams. Inclusion criteria: (1) adults aged 18 years or older; (2) diagnosis of bipolar disorder and in a current depressive episode, defined by Diagnostic Statistic Manual of Mental Disorders, Fifth Edition (DSM-5) (American Psychiatric Association, 2013), determined by a structured assessment using the Mini-International Neuropsychiatric Interview (MINI; Version 7.0.2) (Sheehan et al., 1998); (3) having at least a moderate severity of depressive symptoms as measured by a minimum score of 18 on the Montgomery-Åsberg Depression Rating Scale (MADRS) (Montgomery and Åsberg, 1979); (4) taking a stable dosage of mood-stabilizing medication for a minimum of two weeks or not taking any medication for a minimum of two weeks. Exclusion criteria: (1) symptoms of mania or hypomania as measured by a score of 8 or greater on the Young Mania Rating Scale (YMRS) (Young et al., 1978); (2) any concurrent psychiatric disorders as defined by DSM-5 Axis I or II; (3) having a significant risk of suicide; (4) a history of seizure which resulted in a loss of consciousness; (5) a history of neurological disorder or history of migraines; (7) any exclusion criteria which prevents tDCS administration, including superficial scalp or skin conditions (e.g. psoriasis or eczema), if contact with the scalp is not possible, having metallic implants including intracranial electrodes, surgical clips, shrapnel or pacemaker.

### 2.3. Clinical assessments

Clinical assessments were conducted at baseline, week 2, and week 6, and a follow up assessment was made at month 5 following the initial tDCS session. Assessments were conducted using the following scales: clinician-rated measures of depressive symptoms, MADRS and Hamilton Depression Rating Scale (HDRS-17) (Hamilton, 1960); self-report measure of depressive symptoms: Patient Health Questionnaire-9 (PHQ-9) (Kroenke et al., 2001); clinician-rated measure of anxiety symptoms: Hamilton Anxiety Rating Scale (HAMA) (Hamilton, 1959); clinician-rated measure of manic symptoms, YMRS (Young et al., 1978); self-report measure of disability and impairment: Sheehan Disability Scale (SDS) (Sheehan, 1893); self-report Quality of Life Enjoyment and Satisfaction Questionnaire (Q-LES-Q) (Endicott et al., 1993). Clinical response was defined as an improvement of 50% or greater in MADRS or HAMD score from baseline. Clinical remission was defined as a MADRS score less than 10 and a HAMD score of less than 8. The same researcher was present at each visit and completed ratings for each participant throughout the study as much as possible with clinical supervision from the principal investigator.

### 2.4. Safety, tolerability and acceptability

Safety and tolerability were evaluated by monitoring of adverse events before and after each treatment session using the tDCS Adverse Events Questionnaire (AEQ) (Brunoni et al., 2011). We developed an acceptability questionnaire based on Sekhon et al. (2017) framework model (Woodham et al., 2022) The acceptability questionnaire consisted of five questions were centred on acceptability sub-facets: (1) overall acceptability: ‘How acceptable did you find the tDCS session and how do you feel about the session overall?’; (2) subjective efficacy: ‘How helpful were the tDCS session for improving your depressive symptoms?’; (3) adverse effects: ‘How likely do you think there will be negative side effects from the tDCS session?’; (4) ethical perspectives: ‘How ethical do you think the tDCS session are?’; (5) overall burden: ‘How much effort is required for the tDCS session?’. Responses were assessed on a 7-point Likert style scale along with open-ended responses. Acceptability data were acquired at baseline and week 6. An additional question and four open-ended questions were asked at week 6: (6) retrospective attitude: ‘Would you recommend tDCS to others?’; (7) Positive aspects: ‘What were the most successful parts of the study?’; (8) Negative aspects: ‘What were the least successful parts of the study?’; (9) Possible improvements: ‘How do you think the study could have been improved?’; (10) Further comments: ‘Do you have anything you would like to add, or any further comments?’. Participants completed the questionnaire in a semi-structured interview recorded on video using Microsoft Teams.

### 2.5. Statistical analysis

An intention-to-treat analysis (ITT) was completed including all participants who completed at least one session of tDCS. Last observation carried forward (LOCF) method was used for missing data on clinical assessments. Six repeated-measures ANOVAS were calculated with HDRS-17, MADRS, HAMA, YMRS, PHQ-9 and SDS. The dependent variables were the total scores, and the assessment time-points were the within-subject factor, consisting of three levels: week 0, baseline (t_0_), week 2, after session 10 (t_1_) and week 6, end of treatment period (t_2_). Completers analyses were conducted including participants who completed the minimum number of stimulations and the week 6 study visit. Statistical analyses were conducted using IBM SPSS for Windows version 29.0. All analyses were performed using two-tailed significance values of p=0.05. The Greenhouse-Geisser correction was utilized in cases where Mauchley’s assumption of sphericity was violated. Post-hoc pairwise comparisons were performed with Bonferroni corrections. For the acceptability questionnaire, the median and interquartile range were computed for each response at every time point and the nonparametric Wilcoxon signed-rank test was employed to determine significant differences over time, considering the Likert scale, uncertain difference between anchors, and the limited range of response choices.

## 3. Results

### 3.1. Participants

A total of 44 participants were enrolled (31 women), mean age 47.27 ± 12.94 years. At baseline, mean MADRS and HAMD scores were 24.6 ± 2.64 and 20.0 ± 2.62, respectively. Mean duration of the current depressive episode was 0.95 ± 1.93 years (range 0.3 to 12 years). 97% of participants (n=43) completed the minimum of 15 tDCS sessions, mean number of tDCS sessions 19.6 ±1.9 and 93.2% of participants (n=41) completed the full 6-week course of treatment. 86.3% of participants (n=38) were taking mood-stabilising medication, 2% of participants (n=1) were taking antidepressant medication without mood-stabilising medication, 5 participants were not taking any pharmacological interventions, and 27.3% of participants (n=12) were in psychotherapy (CBT or psychodynamic psychotherapy) in addition to taking medication.

### 3.2. Clinical assessments

For all three time points (weeks 0, 2 and 6), 93% of participants (n=41) completed clinical questionnaires assessments at all time points and were included in the completers analysis. Data was missing from 7% of participants (n=3) at the end of treatment (week 6).

At week 6, mean MADRS score was 8.91 ± 5.56, in which 34 participants (77.3%) showed clinical response and 21 participants (47.7%) achieved clinical remission (Figure. 1). Seven participants (15.9%) showed an early response at week 2, following 10 tDCS sessions, one participant (2.3%) was in remission and mean MADRS score was 16.93 ± 4.82. Repeated-measured analyses demonstrated significant clinical improvements in mean MADRS scores across time points in the ITT (F_(2,86)_= 207.08, p<0.001) (Table. 2) and completers (F_(2,80)_=213.78, p<0.001) analyses (Table. 3). Post hoc tests revealed significant improvements between each of the three time points (p<0.001).

**Figure 1.**
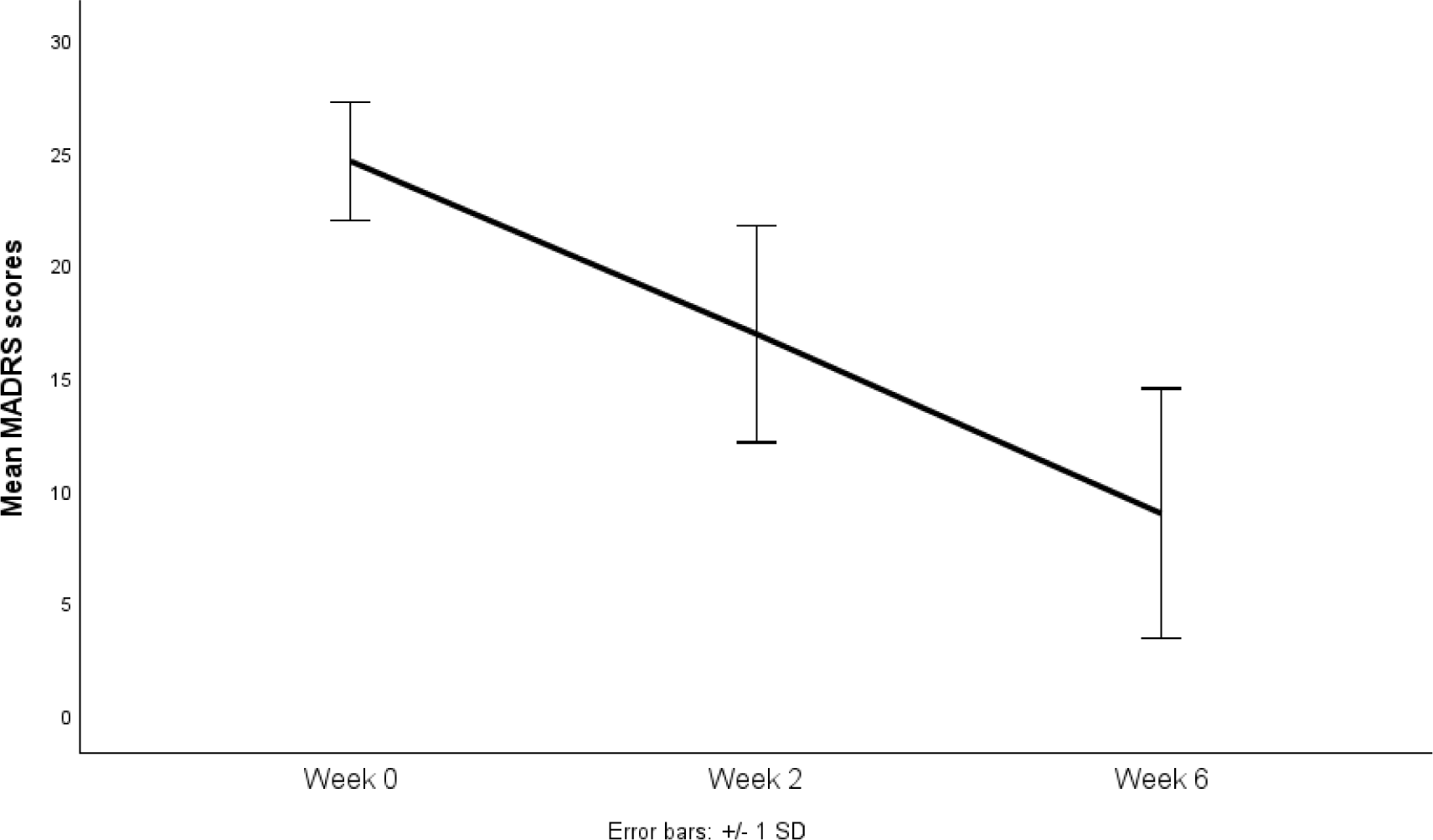
Mean Montgomery-Asberg Depression Rating Scale (MADRS) scores from baseline to week-6 (intention-to-treat analysis). Error bars represent 1 standard deviation.

**Table 1.** Demographic and clinical data at baseline.

|  |  |
| --- | --- |
| Total number (Female) | 44 (31) |
| Mean Age (years) | 47.27 ± 12.89 |
| Age range (years) | 24-76 |
| Age of onset (years) | 27.80 ± 9.28 |
| Years of education | 16.30 ± 2.46 |
| IQ | 100.66 ± 9.29 |
| Duration of illness (years) | 18.98 ± 12.47 |
| Duration current depressive episode (weeks) (range) | 49.55 ± 100.4 |
| Previous number of episodes | 18.16 ± 16.13 |
| Clinical ratings |  |
| MADRS | 24.59 ± 2.64 |
| HDRS-17 | 19.98 ± 2.62 |
| HAMA | 16.55 ± 5.26 |
| YMRS | 2.20 ± 1.49 |
| PHQ-9 | 16.80 ± 4.94 |
| SDS | 20.77 ± 5.87 |
| Treatments during trial |  |
| Taking mood stabilizer and other medications | 38 (86) |
| Taking antidepressant medication only | 1 (2) |
| Taking no medication | 5 (11) |
| Engaged in psychotherapy | 12 (27) |

Categorical variables are presented as number of participants with percentage in parentheses for treatments during trial. Mean values are presented with ' ± ' standard deviation values. MADRS, Montgomery-Åsberg Depression Rating Scale; HDRS-17, Hamilton Depression Rating Scale; HAMA, Hamilton Anxiety Rating Scale; YMRS, Young Mania Rating Scale; PHQ-9, Patient Health Questionnaire-9; SDS, Sheehan Disability Scale.

**Table 2.** Clinical rating scale scores over the course of treatment, intension to treat analysis.

|  | Baseline | Week-2 | Week-6 | F-Value | P-Value |
| --- | --- | --- | --- | --- | --- |
| MADRS | 24.59 ± 2.64 | 16.93 ± 4.82 | 8.77 ± 5.37 | 207.08 | P<0.001 |
| HDRS-17 | 19.98 ± 2.62 | 13.57 ± 4.14 | 6.77 ± 4.74 | 155.90 | P<0.001 |
| HAMA | 16.55 ± 5.26 | 10.43 ± 4.61 | 6.36 ± 4.10 | 82.97 | P<0.001 |
| YMRS | 2.20 ± 1.49 | 1.50 ± 1.15 | 0.80 ± 1.09 | 16.72 | P<0.001 |
| PHQ-9 | 16.80 ± 4.02 | 10.93 ± 4.94 | 6.52 ± 4.69 | 79.83 | P<0.001 |
| SDS | 20.77 ± 5.87 | 16.39 ± 8.06 | 9.93 ± 7.85 | 48.26 | P<0.001 |

Based on intention to treat analysis, using last observation carried forward (n=44). MADRS, Montgomery Asberg Depression Rating Scale; HDRS-17, Hamilton Depression Rating Scale; HAMA, Hamilton Anxiety Rating Scale; Young Mania Rating Scale, YMRS; PHQ-9, Patient Health Questionnaire-9; SDS, Sheehan Disability Scale. Mean values are presented with ' ± ' standard deviation values.

**Table 3.** Clinical rating scale scores over the course of treatment, completers analysis.

|  | Baseline | Week-2 | Week-6 | F-Value | P-Value |
| --- | --- | --- | --- | --- | --- |
| MADRS | 24.68 ± 2.71 | 16.93 ± 4.98 | 8.32 ± 5.27 | 213.78 | P<0.001 |
| HDRS-17 | 20.24 ± 2.41 | 13.63 ± 4.18 | 6.34 ± 4.53 | 177.30 | P<0.001 |
| HAMA | 16.83 ± 5.23 | 10.44 ± 4.65 | 6.07 ± 3.95 | 87.61 | P<0.001 |
| YMRS | 2.27 ± 1.50 | 1.51 ± 1.17 | 0.76 ± 1.09 | 18.07 | P<0.001 |
| PHQ-9 | 16.98 ± 4.07 | 10.85 ± 4.93 | 6.12 ± 4.41 | 85.56 | P<0.001 |
| SDS | 20.83 ± 6.06 | 16.46 ± 8.10 | 9.54 ± 7.74 | 50.61 | P<0.001 |

Data from participants who completed treatment (n=41). MADRS, Montgomery Asberg Depression Rating Scale; HDRS-17, Hamilton Depression Rating Scale; HAMA, Hamilton Anxiety Rating Scale; YMRS, Young Mania Rating Scale; PHQ-9, Patient Health Questionnaire-9; SDS, Sheehan Disability Scale. Mean values are presented with ' ± ' standard deviation values. Demographic data at baseline: female, n=30; mean age 47.93 ± 13.17 years; age range, 24-76 years; age of onset, 27.71 ± 9.45 years; years of education, 16.29 ± 2.50; IQ, 101.24 ± 9.34; duration of illness, 19.53 ± 12.65 years; duration of current depressive episode, 50.68 ± 103.79 weeks; previous number of episodes, 17.36 ± 15.92

HDRS-17 showed a similar pattern of results. Mean HDRS-17 score was 6.77 ± 4.74 at week 6, in which 37 participants (84.1%) showed clinical response and 31 participants (70.5%) achieved clinical remission (Figure. 2). At week 2, 12 participants (27.3%) showed an early response, 3 participants (6.8%) were in remission and mean HDRS-17 score was 13.57 ± 4.14. Significant clinical improvements in mean HDRS-17 scores were evident across time points in ITT (F_(2,86)_=155.9, p<0.001) and completers analyses (F_(2,80)_= 177.30, p<0.001).

**Figure 2.**
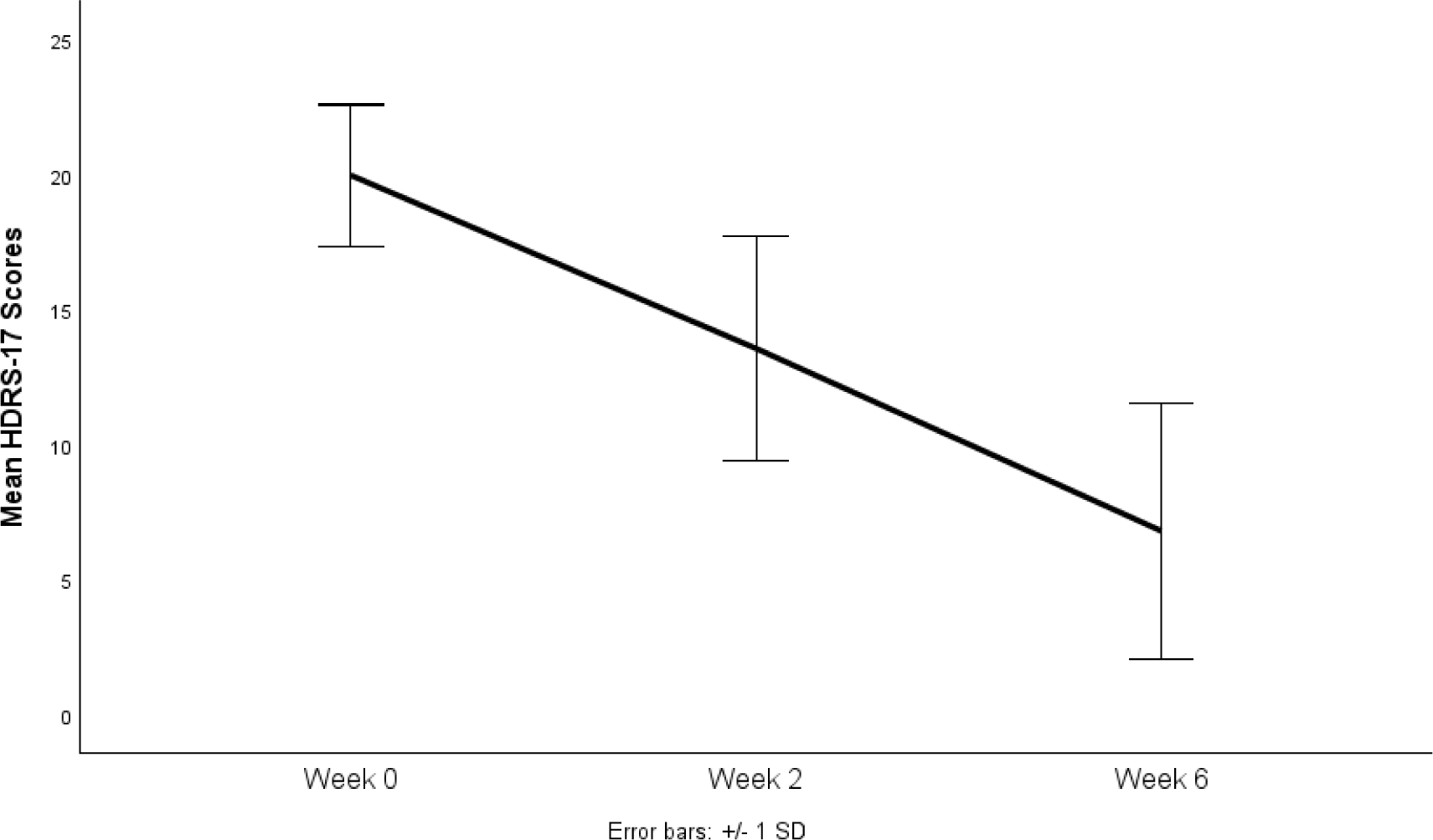
Mean Hamilton Depression Rating Scale (HDRS-17) scores from baseline to week-6 (intention-to-treat analysis). Error bars represent 1 standard deviation.

HAMA, YMRS, PHQ-9 and SDS scores showed significant improvements from baseline and were maintained from week 2 to week 6 (Tables 2 and 3). Mean HAMA score at baseline was 16.6 ± 5.26 (range 9-36), indicating mild to moderate severity of anxiety. Following treatment, the mean score was 6.36 ± 4.10 demonstrating mild anxiety (Figure. 3). Mean YMRS score at baseline was 2.20 ± 1.49 (range 0-7), indicating an overall absence of significant manic or hypomanic symptoms. Following treatment, the mean score decreased to 0.80 ± 1.09 demonstrating a reduction from initial manic or hypomanic symptoms (Figure. 4). Mean PHQ-9 score at baseline was 16.8 ± 4.02, which improved following treatment (mean 6.52 ± 4.69) (Figure. 5). SDS rating of functional impairment was high at baseline (mean 20.77 ± 5.87) and significantly improved at the end of treatment (mean 9.93 ± 7.85) (Figure. 6).

**Figure 3.**
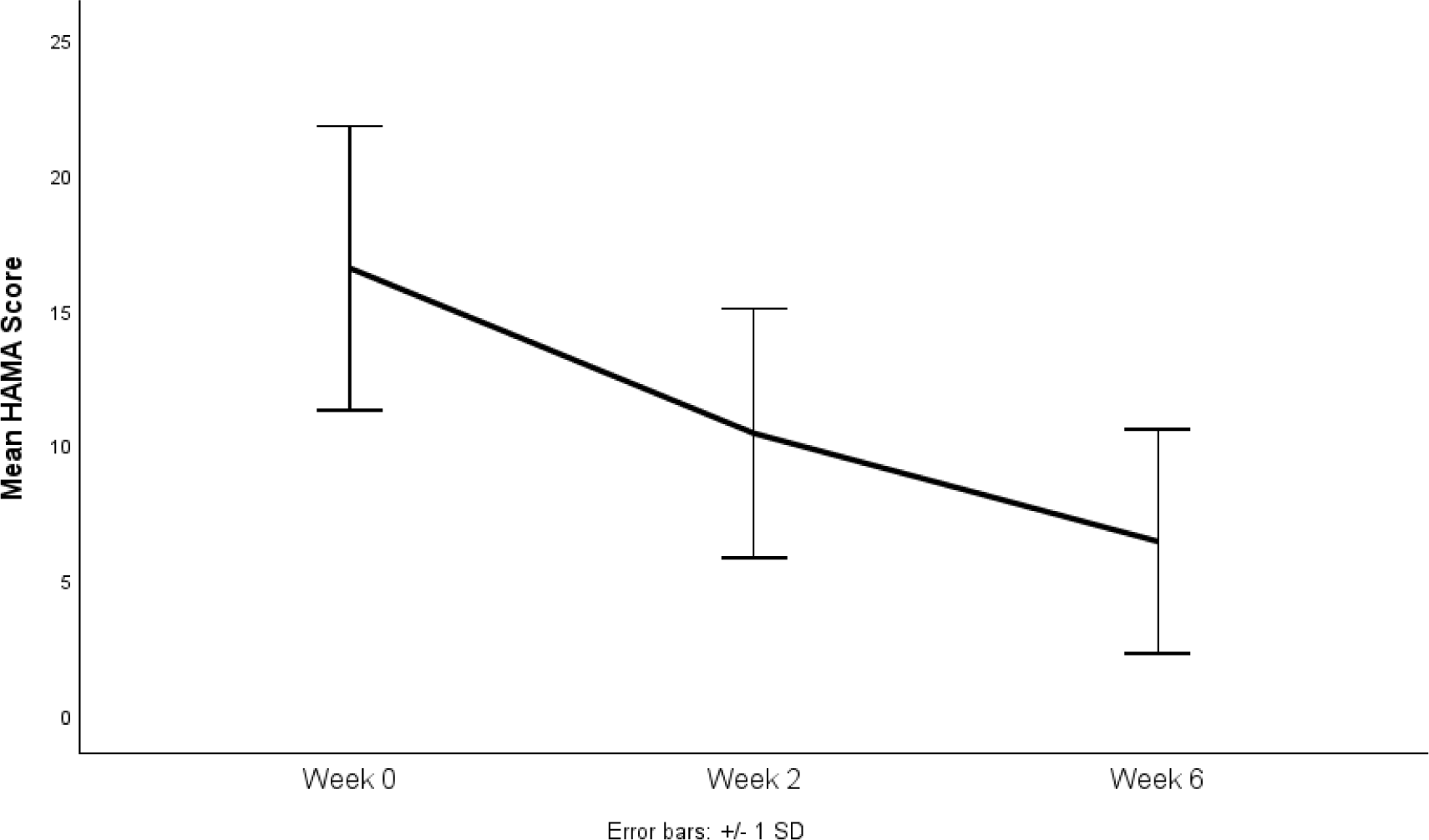
Mean Hamilton Anxiety Scale (HAMA) scores from baseline to week-6 (intention-to-treat analysis). Error bars represent 1 standard deviation.

**Figure 4.**
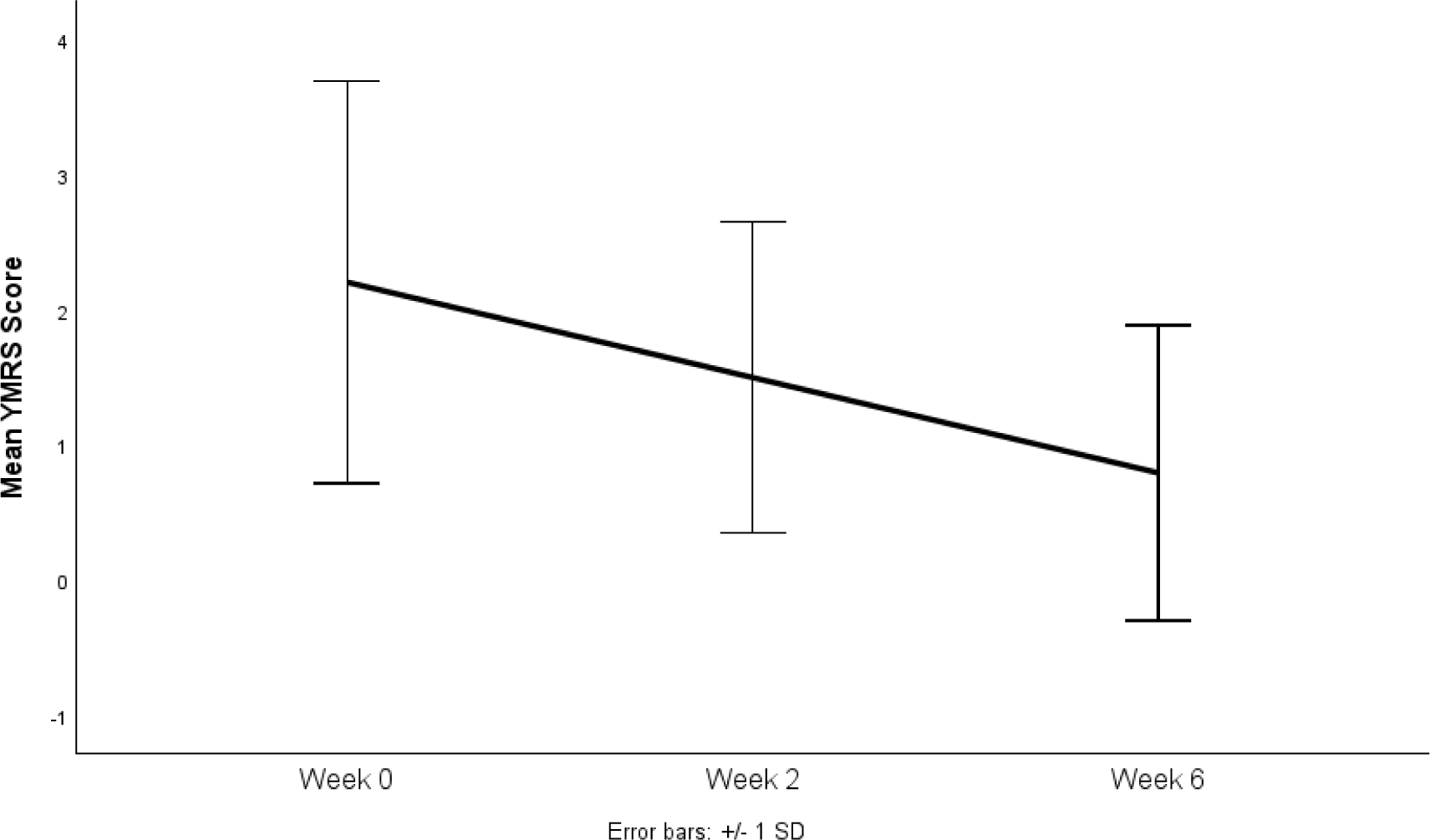
Mean Young Mania Rating Scale (YMRS) scores from baseline to week-6 (intention-to-treat analysis). Error bars represent 1 standard deviation.

**Figure 5.**
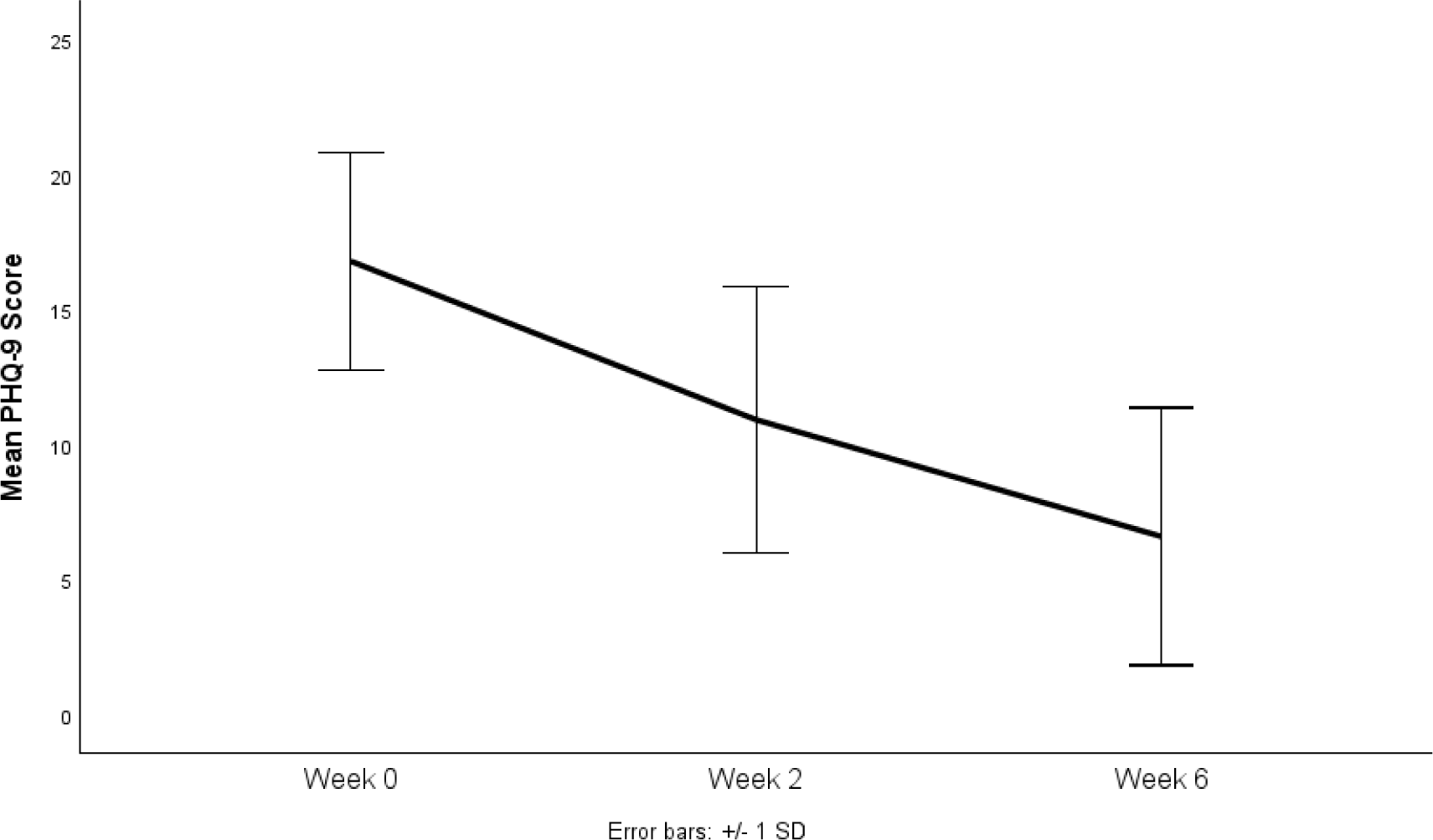
Mean Patient Health Questionnaire-9 (PHQ-9) scores from baseline to week-6 (intention-to-treat analysis). Error bars represent 1 standard deviation.

**Figure 6.**
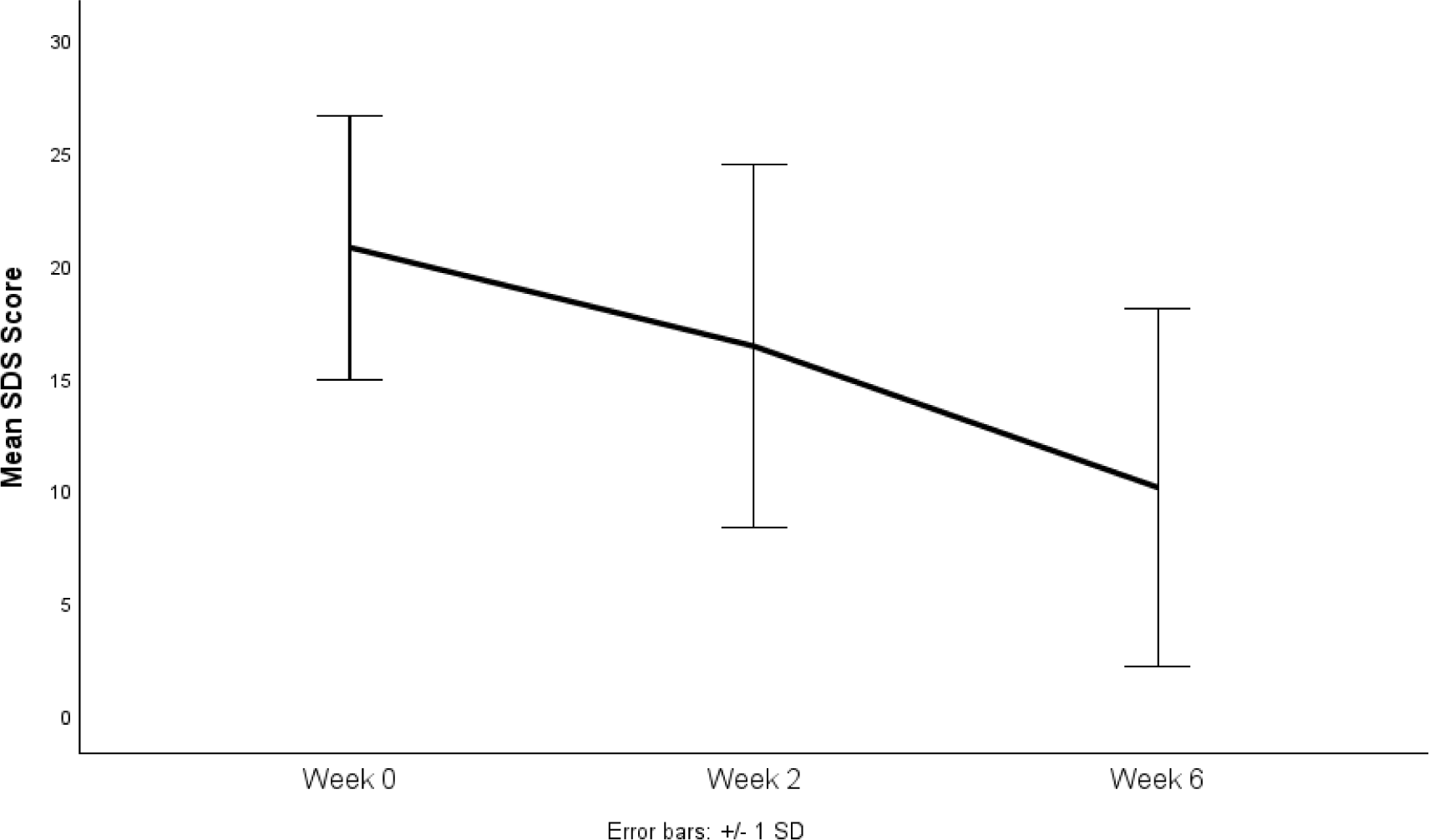
Mean Sheehan Disability Scale (SDS) scores from baseline to week-6 (intention-to-treat analysis). Error bars represent 1 standard deviation.

**Figure 7.**
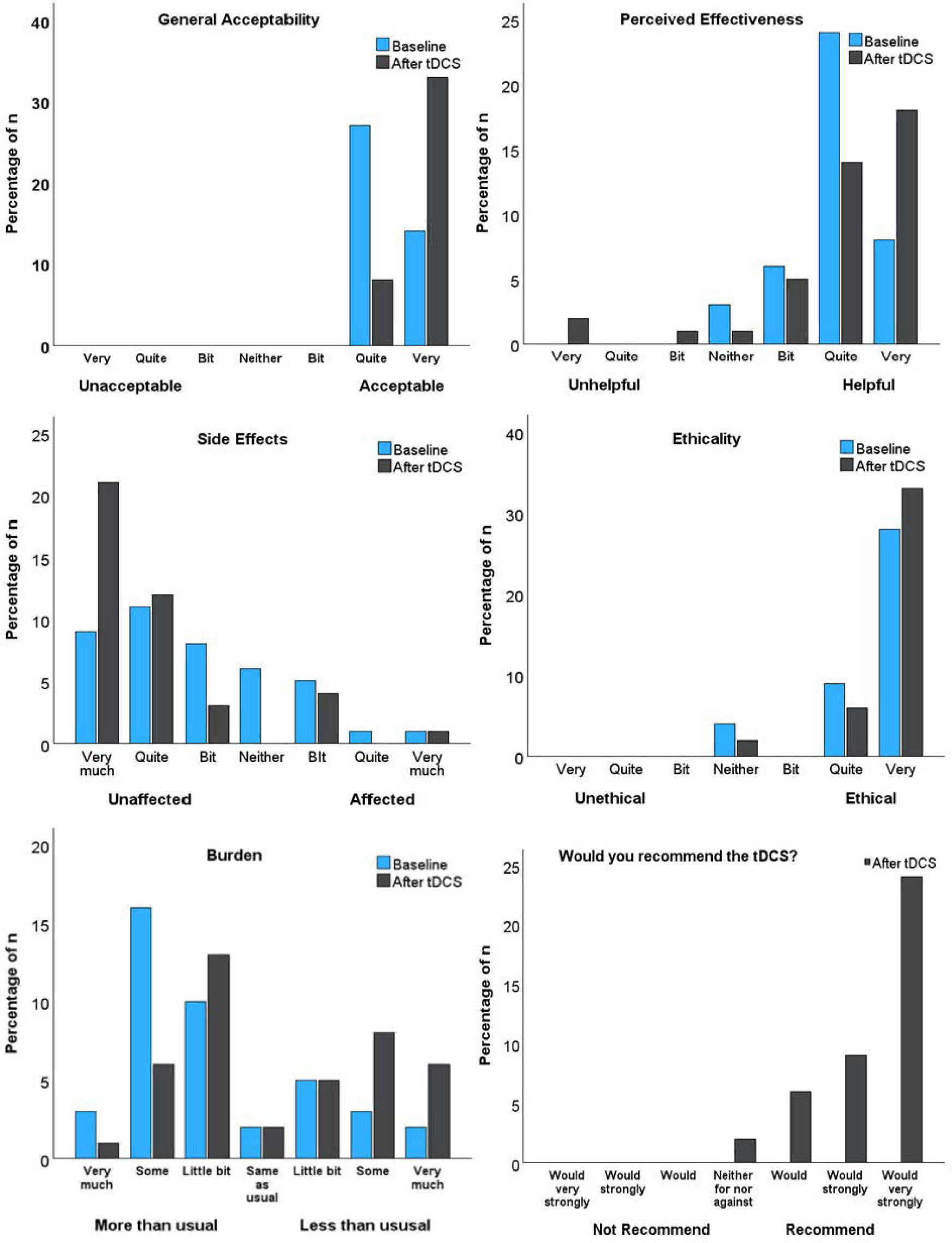
Percentage of participants who endorsed each response in the acceptability questionnaire (completers analysis).

### 3.3. Safety, acceptability, and tolerability

The most common side effects were tingling (83.5%), skin redness (40.6%), itching (29.3%) and burning sensation, (26.5%) (Table 4, supplementary material Table 1). 90.6% of adverse events related to tDCS were rated as mild, 9% were rated as moderate and 0.4% were rated as severe. These included one report each of tingling and burning sensation and two reports each of itching and skin redness.

**Table 4.** Total incidence of side effects out of 860 sessions.

| Side effect | Incidence | Percentage |
| --- | --- | --- |
| Headache | 19 | 2.2% |
| Neck pain | 0 | 0.0% |
| Scalp pain | 24 | 2.8% |
| Tingling | 718 | 83.5% |
| Itching | 252 | 29.3% |
| Burning sensation | 228 | 26.5% |
| Skin redness | 349 | 40.6% |
| Sleepiness | 17 | 2.0% |
| Trouble concentrating | 3 | 0.3% |
| Acute mood change | 6 | 0.7% |
| Other | 18 | 2.1% |
| Tinnitus | 1 | 0.1% |
| Pressure on right eye | 1 | 0.1% |
| Vibration | 3 | 0.3% |
| Improved concentration | 1 | 0.1% |
| stinging | 1 | 0.1% |
| Sore feeling | 2 | 0.2% |
| vivid dreams | 2 | 0.2% |
| Dizziness | 2 | 0.2% |
| Bruise | 1 | 0.1% |
| Dry skin | 3 | 0.3% |
| Throbbing in left eye | 1 | 0.1% |

Adverse events were recorded using the tDCS Adverse Events Questionnaire (Brunoni et al., 2011). An adverse event was present if the participant rated that it was at least remotely possible that it was associated with the intervention.

There was a significant increase in endorsement of acceptability as being “quite acceptable” at baseline and “very acceptable” post treatment (t_(0)_ Mdn = 6, IQR = 1; t_(2)_ Mdn = 7, IQR = 0) (*Z* = -4.15, *p* < 0.001). Ratings for perceived effectiveness was endorsed as being “Quite helpful” at baseline and post treatment with no significant change over time (t_(0)_ Mdn = 6, IQR = 0; t_(2)_ Mdn = 6, IQR = 1) (*Z* = -0.95, *p* = 0.34). Ethicality remained high at “very ethical” with no significant changes over time (t_(0)_ Mdn = 7, IQR = 1; t_(2)_ Mdn = 7, IQR = 0) (*Z* = -1.43, *p* = 0.15). The impact of side-effects showed a significant decrease from being “a bit unaffected” at baseline to being “very much unaffected” post-treatment (t_(0)_ Mdn = 3, IQR = 2; t_(2)_ Mdn = 1, IQR = 1) (*Z* = -2.77, *p* = 0.006). There was also a significant decrease in the perceived amount of effort required to remain consistent which improved from “little bit more effort than usual” at baseline to “about the same effort as usual” post treatment (t_(0)_ Mdn = 3, IQR = 2.5; t_(2)_ Mdn = 4, IQR = 3) (*Z* = -3.70, *p* < 0.001). At week 6 participants “would very strongly recommend” tDCS to others (t_(2)_ Mdn = 7, IQR = 1) (Figure.7).

## 4. Discussion

The present 6-week course of home-based tDCS with real-time supervision was associated with significant clinical improvements in bipolar depression, high rates of clinical response and remission, high treatment acceptability, and mild adverse effects. Depressive rating n scores at each time-point showed a consistent decrease in both clinician-rated and self-rated measures. Moreover, anxiety symptoms and disability measures were significantly improved at the end of treatment.

Adverse events related to tDCS were mild over 90% of the time and transient. The most common side effects were tingling, skin redness, itching and burning sensation, which are typical with tDCS (Brunoni et al., 2011). There were no serious adverse events associated with the device, nor were there any instances of treatment-emergent affective switching. Remote real-time supervision during stimulation allowed for close monitoring of adverse events and ensuring that the device was used correctly. Monitoring of side effects with an online daily report has also been effective in a home-based trial (Alonzo et al., 2019) as well as periodic monitoring visits in two large home-based trials (Borrione et al., 2024; Woodham et al., 2023). Safety reporting for home-based tDCS treatments is an important consideration as reports of skin burns at the electrode site have been reported (Kumpf et al., 2023; Woodham et al., 2023), which can occur with insufficient moistening with conductive saline solution (Kortteenniemi et al., 2019) or application of tap water to moisten sponges (Frank et al., 2010; Palm et al., 2008). If participants encounter potential challenges in managing side effects independently, it could lead to the exacerbation of adverse events and eventual discontinuation of treatment.

In bipolar depression, Lee et al. (2022) recently reported an RCT of daily home-based active or sham tDCS as an adjunct treatment to mood-stabilising medication. No significant differences in depressive symptoms were found between groups after 6-weeks of treatment. However, the trial had included five in person clinical visits, and only 59.3% of participants completed the full course of treatment, and missed in person assessments were an exclusion criterion. In the present study, the protocol was fully remote with real-time visits during each session and the discontinuation rate was 6.8%.

While there is no definitive consensus regarding optimal scheduling or dosage, the present study used parameters established through meta-analyses, which demonstrated highly effective treatment outcomes using a minimum of 20 sessions, lasting 30 minutes each, with electrical current set at 2mA (Brunoni et al., 2016; Mutz et al., 2019, 2018; Woodham et al., 2022), but increased sessions frequency have been correlated improved clinical outcomes (Moffa et al., 2020). As far as we are aware, the present study is the first to employ remote supervision for home-based tDCS for bipolar depression. A significant advantage of employing a home-based tDCS protocols is that participants had the autonomy to schedule their sessions according to their preferences, thereby enabling them to maintain a consistent regimen at a convenient time. This may have contributed to the low attrition rate which was notably lower than clinic-based tDCS protocols, in which the attrition rate might reach 10.1% (Brunoni et al., 2016; Mutz et al., 2019). Furthermore, the present study found significant improvements in disability and functional outcomes, which has been observed with TMS (Tavares et al., 2017).

Limitations of the study include the absence of a sham tDCS treatment arm as all participants received active tDCS with an open-label design. The provision of real-time supervision for each session likely played a role in the improvement of depressive symptoms (Papoutsi and Fu, 2021). Furthermore, the study did not control for the types of medication. Participants were required to maintain a stable dosage of mood stabilizing medication for at least two weeks or abstain from medication for the same duration. Mood stabilizers such as lithium and lamotrigine, exert their effects through the modulation of cortical excitability, a mechanism shared with voltage-gated sodium channels (Lee et al., 2022). A reduction in cortical excitability could be associated with a reduction in tDCS efficacy (Romero Lauro et al., 2014). Variations in head sizes, individual anatomical characteristics, and device placement among users may have resulted in different configurations of electrical field density with the brain. Individual differences in tDCS effects may arise partially due to discrepancies in electric fields. The tDCS device used in the current study underwent electric field modelling, which indicates its targeting of areas implicated in the pathophysiology of MDD within the prefrontal cortex. However, differences in the device positioning may have influenced the intensity of the electric field and subsequently impacted treatment outcomes.

In summary, home-based tDCS with real-time remote supervision was associated with significant improvements in depressive symptoms in individuals with bipolar depression of moderate to severe severity. The present study demonstrated high levels of acceptability, tolerability and safety for home-based tDCS in bipolar depression.

## Data Availability

Anonymised data will be made available on request to the corresponding author.

