## Supplementary Material for "Home-based transcranial direct current stimulation in bipolar depression: an open-label treatment study of clinical outcomes, acceptability and adverse events"

**Supplementary Materials**

**Supplementary Material Figure 1**. Diagram showing study design and progress of participants through the trial.

Interview assessment for eligibility (*n*=66)

Excluded (*n*=22)

- No bipolar diagnosis: 2
- Informed consent not provided: 1
- Significant suicide ideation: 5
- Concurrent mental health disorder: 5
- Migraine: 2
- Stroke: 1
- Declined to participate: 4
- Lives outside England and Wales: 1
- Not in current depressive episode: 1

Enrolled participants (*n*=44)

Discontinued participation (*n*=1)

Did not attend final visit (n=2)

6-week primary endpoint analysis (*n*=41)

Long term 3-month follow up (*n*=32)

**Supplementary Materials Table 1.** Incidence of adverse events at each study visit.

|  | V1 | V2 | V3 | V4 | V5 | V6 | V7 | V8 | V9 | V10 | V11 |
| --- | --- | --- | --- | --- | --- | --- | --- | --- | --- | --- | --- |
| Total no. of participants | 44 | 44 | 43 | 40 | 35 | 41 | 44 | 42 | 42 | 44 | 42 |
| **Side effect** |  |  |  |  |  |  |  |  |  |  |  |
| Headache | 3 (6.8) | 1 (2.3) | 3 (7) | 1 (2.5) | 1 (2.9) | 0 (0) | 0 (0) | 0 (0) | 1 (2.4) | 0 (0) | 2 (4.8) |
| Neck pain | 0 (0) | 0 (0) | 0 (0) | 0 (0) | 0 (0) | 0 (0) | 0 (0) | 0 (0) | 0 (0) | 0 (0) | 0 (0) |
| Scalp pain | 5 (11.4) | 1 (2.3) | 1 (2.3) | 1 (2.5) | 1 (2.5) | 1 (2.4) | 2 (4.5) | 2 (4.8) | 1 (2.4) | 1 (2.3) | 1 (2.4) |
| Tingling | 42 (95.5) | 41 (93.2) | 39 (90.7) | 38 (95) | 31 (88.6) | 35 (85.4) | 38 (86.4) | 34 (81) | 37 (88.1) | 35 (79.5) | 34 (81) |
| Itching | 20 (45.5) | 18 (40.9) | 19 (44.2) | 15 (37.5) | 15 (42.9) | 15 (36.6) | 14 (31.8) | 13 (31) | 12 (28.6) | 11 (25) | 10 (23.8) |
| Burning sensation | 15 (34.1) | 18 (40.9) | 16 (37.2) | 13 (32.5) | 11 (31.4) | 12 (29.9) | 15 (34.1) | 14 (33.3) | 18 (42.9) | 14 (31.8) | 12 (28.6) |
| Skin redness | 19 (43.2) | 22 (50) | 23 (53.5) | 21 (52.5) | 13 (37.1) | 18 (43.9) | 21 (47.7) | 20 (0) | 17 (40.5) | 19 (43.2) | 16 (38.1) |
| Sleepiness | 3 (6.8) | 1 (2.3) | 1 (2.3) | 1 (2.5) | 0 (0) | 1 (2.4) | 1 (2.3) | 2 (4.8) | 1 (2.4) | 2 (4.5) | 2 (4.8) |
| Trouble concentrating | 0 (0) | 1 (2.3) | 0 (0) | 0 (0) | 0 (0) | 0 (0) | 0 (0) | 0 (0) | 0 (0) | 0 (0) | 0 (0) |
| Acute mood change | 3 (6.8) | 2 (4.5) | 0 (0) | 1 (2.5) | 0 (0) | 0 (0) | 0 (0) | 0 (0) | 0 (0) | 0 (0) | 0 (0) |
| Other | 2 (4.5) | 4 (6.8) | 1 (2.3) | 2 (5) | 2 (5.7) | 0 (0) | 0 (0) | 0 (0) | 1 (2.4) | 0 (0) | 0 (0) |
| Tinnitus | 0 (0) | 1 (2.3) | 0 (0) | 0 (0) | 0 (0) | 0 (0) | 0 (0) | 0 (0) | 0 (0) | 0 (0) | 0 (0) |
| Pressure on right eye | 0 (0) | 1 (2.3) | 0 (0) | 0 (0) | 0 (0) | 0 (0) | 0 (0) | 0 (0) | 0 (0) | 0 (0) | 0 (0) |
| Vibration | 1 (2.3) | 1 (2.3) | 1 (2.3) | 0 (0) | 0 (0) | 0 (0) | 0 (0) | 0 (0) | 0 (0) | 0 (0) | 0 (0) |
| Improved concentration | 0 (0) | 0 (0) | 0 (0) | 1 (2.5) | 0 (0) | 0 (0) | 0 (0) | 0 (0) | 0 (0) | 0 (0) | 0 (0) |
| stinging | 0 (0) | 0 (0) | 0 (0) | 1 (2.5) | 0 (0) | 0 (0) | 0 (0) | 0 (0) | 0 (0) | 0 (0) | 0 (0) |
| Sore feeling | 0 (0) | 0 (0) | 0 (0) | 0 (0) | 2 (5.7) | 0 (0) | 0 (0) | 0 (0) | 0 (0) | 0 (0) | 0 (0) |
| Vivid dreams | 0 (0) | 1 (2.3) | 0 (0) | 0 (0) | 0 (0) | 0 (0) | 0 (0) | 0 (0) | 1 (2.4) | 0 (0) | 0 (0) |
| Dizziness | 1 (2.3) | 0 (0) | 0 (0) | 0 (0) | 0 (0) | 0 (0) | 0 (0) | 0 (0) | 0 (0) | 0 (0) | 0 (0) |
| Bruise | 0 (0) | 0 (0) | 0 (0) | 0 (0) | 0 (0) | 0 (0) | 0 (0) | 0 (0) | 0 (0) | 0 (0) | 0 (0) |
| Dry skin | 0 (0) | 0 (0) | 0 (0) | 0 (0) | 0 (0) | 0 (0) | 0 (0) | 0 (0) | 0 (0) | 0 (0) | 0 (0) |
| Throbbing in left eye | 0 (0) | 0 (0) | 0 (0) | 0 (0) | 0 (0) | 0 (0) | 0 (0) | 0 (0) | 0 (0) | 0 (0) | 0 (0) |

|  | V12 | V13 | V14 | V15 | V16 | V17 | V18 | V19 | V20 | V21 |
| --- | --- | --- | --- | --- | --- | --- | --- | --- | --- | --- |
| Total no. of participants | 40 | 41 | 40 | 37 | 42 | 39 | 40 | 40 | 40 | 40 |
| **Side effect** |  |  |  |  |  |  |  |  |  |  |
| Headache | 0 (0) | 0 (0) | 1 (2.5) | 0 (0) | 2 (4.8) | 1 (2.6) | 2 (5) | 0 (0) | 0 (0) | 1 (2.5) |
| Neck pain | 0 (0) | 0 (0) | 0 (0) | 0 (0) | 0 (0) | 0 (0) | 0 (0) | 0 (0) | 0 (0) | 0 (0) |
| Scalp pain | 1 (2.5) | 1 (2.4) | 1 (2.5) | 1 (2.7) | 1 (2.4) | 0 (0) | 1 (2.5) | 0 (0) | 1 (2.5) | 0 (0) |
| Tingling | 34 (85) | 33 (80.5) | 34 (85) | 30 (0) | 33 (78.6) | 31 (79.5) | 30 (75) | 31 (77.5) | 30 (75) | 28 (70) |
| Itching | 12 (30) | 11 (26.8) | 11 (27.5) | 12 (32.4) | 12 (28.6) | 7 (17.9) | 7 (17.5) | 6 (15) | 6 (15) | 6 (15) |
| Burning sensation | 9 (22.5) | 9 (22) | 8 (22) | 6 (16.2) | 6 (14.3) | 5 (12.8) | 8 (20) | 6 (15) | 7 (17.5) | 6 (15) |
| Skin redness | 16 (40) | 16 (39) | 15 (37.5) | 13 (35.1) | 16 (38.1) | 12 (30.8) | 14 (35) | 15 (37.5) | 12 (30) | 11 (27.5) |
| Sleepiness | 0 (0) | 1 (2.4) | 0 (0) | 0 (0) | 0 (0) | 0 (0) | 1 (2.5) | 0 (0) | 0 (0) | 0 (0) |
| Trouble concentrating | 0 (0) | 1 (2.4) | 1 (2.5) | 0 (0) | 0 (0) | 0 (0) | 0 (0) | 0 (0) | 0 (0) | 0 (0) |
| Acute mood change | 0 (0) | 0 (0) | 0 (0) | 0 (0) | 0 (0) | 0 (0) | 0 (0) | 0 (0) | 0 (0) | 0 (0) |
| Other | 1 (2.5) | 0 (0) | 1 (2.5) | 2 (5.4) | 0 (0) | 1 (2.6) | 0 (0) | 0 (0) | 1 (2.5) | 0 (0) |
| Tinnitus | 0 (0) | 0 (0) | 0 (0) | 0 (0) | 0 (0) | 0 (0) | 0 (0) | 0 (0) | 0 (0) | 0 (0) |
| Pressure on right eye | 0 (0) | 0 (0) | 0 (0) | 0 (0) | 0 (0) | 0 (0) | 0 (0) | 0 (0) | 0 (0) | 0 (0) |
| Vibration | 0 (0) | 0 (0) | 0 (0) | 0 (0) | 0 (0) | 0 (0) | 0 (0) | 0 (0) | 0 (0) | 0 (0) |
| Improved concentration | 0 (0) | 0 (0) | 0 (0) | 0 (0) | 0 (0) | 0 (0) | 0 (0) | 0 (0) | 0 (0) | 0 (0) |
| Stinging | 0 (0) | 0 (0) | 0 (0) | 0 (0) | 0 (0) | 0 (0) | 0 (0) | 0 (0) | 0 (0) | 0 (0) |
| Sore feeling | 0 (0) | 0 (0) | 0 (0) | 0 (0) | 0 (0) | 0 (0) | 0 (0) | 0 (0) | 0 (0) | 0 (0) |
| Vivid dreams | 0 (0) | 0 (0) | 0 (0) | 0 (0) | 0 (0) | 0 (0) | 0 (0) | 0 (0) | 0 (0) | 0 (0) |
| Dizziness | 1 (2.5) | 0 (0) | 0 (0) | 0 (0) | 0 (0) | 0 (0) | 0 (0) | 0 (0) | 0 (0) | 0 (0) |
| Bruise | 0 (0) | 0 (0) | 0 (0) | 1 (2.7) | 0 (0) | 0 (0) | 0 (0) | 0 (0) | 0 (0) | 0 (0) |
| Dry skin | 0 (0) | 0 (0) | 1 (2.5) | 1 (2.7) | 0 (0) | 1 (2.6) | 0 (0) | 0 (0) | 0 (0) | 0 (0) |
| Throbbing in left eye | 0 (0) | 0 (0) | 0 (0) | 0 (0) | 0 (0) | 0 (0) | 0 (0) | 0 (0) | 1 (2.5) | 0 (0) |
| Adverse events were recorded using the tDCS Adverse Events Questionnaire (Brunoni et al., 2011). An adverse event was present if the participant rated that it was at least remotely possible that it was associated with the intervention. The number of participants is presented with the percentage in parenthesis. Participant reported throbbing in left eye for the last 2 minutes of stimulation. Reports are described as presented by participants. | | | | | | | | | | |
